# Clinical and Virological Characteristics of Hospitalized COVID-19 Patients in a German Tertiary Care Center during the First Wave of the SARS-CoV-2 Pandemic

**DOI:** 10.1101/2020.12.12.20247726

**Authors:** Charlotte Thibeault, Barbara Mühlemann, Elisa T. Helbig, Mirja Mittermaier, Tilman Lingscheid, Pinkus Tober-Lau, Lil A. Meyer-Arndt, Leonie Meiners, Paula Stubbemann, Sascha S. Haenel, Laure Bosquillon de Jarcy, Lena Lippert, Moritz Pfeiffer, Miriam S. Stegemann, Robert Roehle, Janine Wiebach, Stefan Hippenstiel, Thomas Zoller, Holger Müller-Redetzky, Alexander Uhrig, Felix Balzer, Christof von Kalle, Norbert Suttorp, Terry C. Jones, Christian Drosten, Martin Witzenrath, Leif E. Sander, Pa-COVID Study Group, Victor M. Corman, Florian Kurth

## Abstract

**Background:** Adequate patient allocation is pivotal for optimal resource management in strained healthcare systems, and requires detailed knowledge of clinical and virological disease trajectories.

**Methods:** A cohort of 168 hospitalized adult COVID-19 patients enrolled in a prospective observational study at a large European tertiary care center was analyzed.

**Results:** Forty-four percent (71/161) of patients required invasive mechanical ventilation (IMV). Shorter duration of symptoms before admission (aOR 1.22 per day less, 95%CI 1.10-1.37, p<0.01), age 60-69 as compared to 18-59 years (aOR 4.33, 95%CI 1.07-20.10, p=0.04), and history of hypertension (aOR 5.55, 95%CI 2.00-16.82, p<0.01) were associated with need for IMV. Patients on IMV had higher maximal concentrations, slower decline rates, and longer shedding of SARS-CoV-2 than non-IMV patients (33 days, IQR 26-46.75, vs 18 days, IQR 16-46.75, respectively, p<0.01). Median duration of hospitalization was 9 days (IQR 6-15.5) for non-IMV and 49.5 days (IQR 36.8-82.5) for IMV-patients.

**Conclusion:** Our results indicate a short duration of symptoms before admission as a risk factor for severe disease and different viral load kinetics in severely affected patients.

## Background

The ongoing Severe Acute Respiratory Syndrome Coronavirus 2 (SARS-CoV-2) pandemic places an unprecedented burden on healthcare systems worldwide. Host factors predictive of severe clinical course and adverse outcome in patients with Coronavirus disease 2019 (COVID-19) include older age, male gender, and pre-existing chronic comorbidities [1-6]. Several risk scores containing clinical characteristics, laboratory assessments, and biomarkers have been proposed for improved patient management and resource allocation [5, 7].

Reported proportions of hospitalized patients requiring invasive mechanical ventilation (IMV) vary considerably between 2.3% [8] and 23.6% [2]. In-hospital case fatality rates of 20-30% have been described in China, Germany, Italy, the UK, and the US [1, 6, 9-11]. In Germany, shortages of inpatient beds and intensive care unit (ICU) capacity were largely avoided during the first pandemic wave [12], contributing to a comparatively low overall case fatality rate [1, 13].

Here, we report clinical characteristics, laboratory and virological parameters, clinical course, and outcome of 168 COVID-19 patients included in a prospective observational cohort study conducted at Charité – Universitätsmedizin Berlin, Germany. The study was designed for deep clinical, molecular, and immunological phenotyping of COVID-19 [14-18].

The data reflect the situation in a tertiary care referral center for the treatment of patients with acute respiratory distress syndrome (ARDS), including veno-venous extracorporeal membrane oxygenation (vvECMO) therapy at an associated certified weaning center during the first months of the COVID-19 pandemic, before treatment with dexamethasone became standard of care [19]. Specific aims of this work were to identify risk factors associated with need for IMV, to analyze viral kinetics in patients with and without IMV, and to provide a comprehensive description of clinical course and outcome.

## Methods

### Study cohort and data collection

Data collection was performed within the Pa-COVID-19 study, a prospective observational cohort study conducted at Charité – Universitätsmedizin Berlin, as described [14]. Adult patients admitted between March 1^st^ and June 30^th^, 2020, with PCR-confirmed SARS-CoV-2 infection were included if patients or their legal representatives gave informed consent. We recorded epidemiological and demographic data, medical history, history of present illness, symptoms, clinical course, treatment, and outcomes upon enrolment and longitudinally during hospitalization. The study was approved by the ethics committee of Charité – Universitätsmedizin Berlin (EA2/066/20), conducted according to the Declaration of Helsinki and Good Clinical Practice principles (ICH 1996) and is registered in the German and WHO international clinical trials registry (DRKS00021688) [14].

Comorbidities were classified using the Charlson comorbidity Index (CCI) [20]. ARDS was defined according to the Berlin definition of ARDS [21]. Sequential Organ Failure Assessment (SOFA) score [22] was calculated from data recorded in the ICU data management system. The following predefined events were assessed in all patients: 1) Sepsis (defined according to sepsis-3 criteria [23], 2) venous thromboembolic events (VTE; pulmonary embolism or deep vein thrombosis), 3) neurological events (hemorrhagic/ischemic stroke, delirium, critical illness polyneuropathy (CIP), critical illness myopathy (CIM), epileptic seizure, meningitis and encephalitis). Treatment was unaffected by participation in the study. Patient allocation was performed according to structured regional processes [24] and management of critically ill patients following national and international recommendations [25, 26].

All laboratory assessments were carried out in accredited laboratories at Charité – Universitätsmedizin Berlin. SARS-CoV-2 viral concentration was measured in respiratory samples (naso-or oropharyngeal swabs) by real-time RT-PCR (32). Viral concentration is given as log_10_ genome copies per swab or initial 1 mL sampling buffer.

### Statistical analyses

Distribution of continuous variables was summarized by median and interquartile range (IQR) values or mean and standard deviation (sd), as appropriate. The differences of continuous variables between groups were examined by Welch’s t-test or, in absence of normal distribution, by Mann-Whitney *U* test. Categorical variables were compared using chi-square tests. For all analyses, complete cases were used for the respective evaluation.

We conducted a multiple logistic regression with “need for invasive mechanical ventilation” as a binary dependent variable and age, BMI, hypertension, diabetes, and time between symptom onset and admission to hospital as independent variables. The covariates were chosen taking into account current evidence, results from univariate testing, and sample size. We performed univariate tests regarding all available patient factors and association with organ support treatment, complications, and outcome. Patients with therapy limitations (Do Not Intubate (DNI) or Do Not Resuscitate (DNR) orders) at the respective time point were excluded for comparison between non-IMV and IMV patients, and for analyses of course-of-organ support and mortality. Non-survivors were excluded from comparison between short- (< 15 days) or long-term (≥ 15 days) IMV. For analysis of SOFA score, the highest score per day was included for calculation of means. The neurological component of the SOFA score was not taken into consideration due to patient sedation. Patients who died or were transferred to other centers were excluded from calculation of length of hospital stay. For analyses of routine laboratory parameters within 72 hours of first admission, first-available parameters were included.

We compared viral concentration between non-IMV and IMV patients, and regressed viral concentration on the duration from symptom onset to admission using both the first positive RT-PCR result and the RT-PCR with the highest viral concentration. For the calculation of viral concentration decline, we estimated the slope parameter from a linear regression of at least four viral concentration measurements over time for each patient. If available, the first of at least two final negative RT-PCR results was included, in which case the viral concentration of the negative RT-PCR was set to 2.0 in accordance with the RT-PCR limit of detection and sample dilution factor of ∼20 [27, 28]. We calculated shedding duration as the time from symptom onset to the date of the first of at least two final negative RT-PCR results.

Analyses were conducted with R (version 3.6.1) [29], JMP (version 14.2.0) [30], and statsmodels (version 0.12.0) in Python 3.7.9 [31]. A p-value <0.05 indicates statistical significance, although all results have to be considered as non-confirmatory. For this reason, no adjustment for multiple testing was done.

## Results

### Baseline characteristics

Between March 1^st^ and June 30^th^ 2020, a total of 347 adult patients with COVID-19 were hospitalized at Charité – Universitätsmedizin Berlin. This analysis includes 168 adult patients who consented to participation in the prospective observational study (**Figure 1**). Sixty-five percent (110/168) were directly admitted to our center, whereas 29.8% (50/168) were referred due to ARDS or other conditions requiring tertiary care. Four percent (7/168) were hospitalized for other reasons and coincidentally diagnosed with SARS-CoV-2 infection during routine screening, and one patient (0.6%) was admitted due to a late complication of COVID-19.

**Figure 1:**
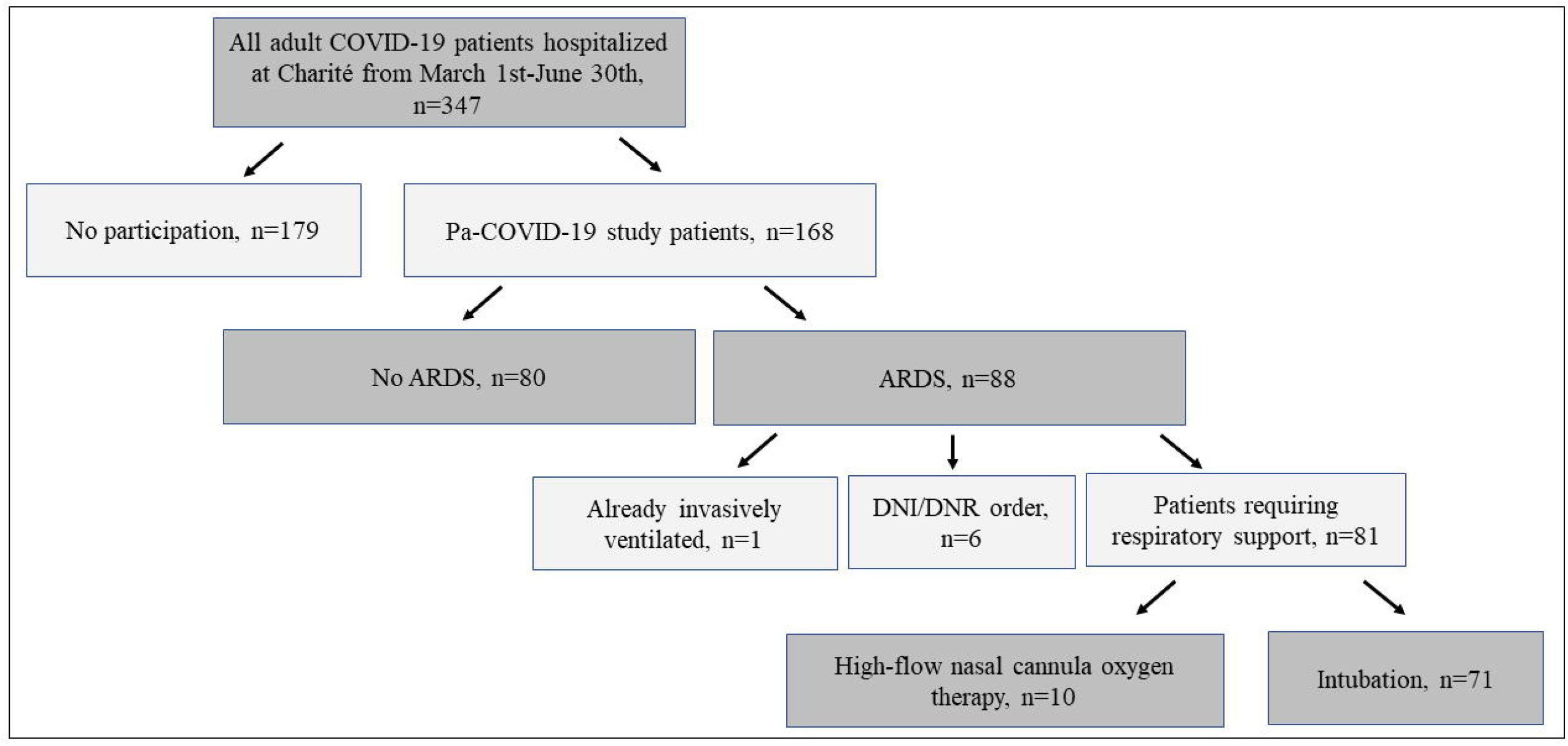
Study cohort flowchart. A total of 347 adult patients were hospitalized with COVID-19 during the study period from March 1^st^ until June 30^th^ at Charité-Universitätsmedizin Berlin. Of these, 168 patients could be enrolled in the prospective observational study, whereas 179 denied. Among the included patients, 88 had acute respiratory distress syndrome (ARDS). One patient with ARDS was already invasively ventilated and six of them had DNI/DNR (do not intubate/do not resuscitate) orders in place, resulting in 81 patients requiring respiratory support. Of those, 71 patients were intubated and ten required only high-flow nasal cannula oxygen therapy.

Baseline characteristics are shown in **<u>Table 1</u>**. Median patient age was 61 years (IQR 49.3-72), and 66.1% (111/168) were male. Median CCI was 3 (IQR 1-4). Most prevalent comorbidities were hypertension (53.6%, 90/168), diabetes (19.6%, 33/168), and chronic pulmonary disease (16.7%, 28/168). Median time from symptom onset until diagnosis was 4 days (IQR 1-7), and from symptom onset until admission to hospital was 6 days (IQR 3-10).

### ARDS and organ support treatment

Fifty-two percent (88/168) of patients developed ARDS. One of them was already dependent on long-term intermittent invasive ventilation and six had DNI orders in place. Of the remaining 81 patients with ARDS, 87.6% (71/81) required IMV whereas 12.3% (10/81) could be managed with HFNC oxygen only (**Figure 1**). Median time from hospital admission to intubation was 2 days (IQR 0-4). Among all patients without therapeutic limitations, 44.1% (71/161) required IMV, 9.9% (16/161) could be treated with HFNC oxygen only, 24.2% (39/161) required oxygen via nasal prongs, and 22% (35/161) were not in need of supplemental oxygen.

Age between 60 and 69 years as compared to 18 to 59 years (adjusted OR 4.33, 95% CI 1.07-20.10, p=0.05) and pre-existing hypertension (adjusted OR 5.55, 95% CI 2.00-16.82, p<0.01) were independent risk factors for IMV requirement in multivariable analysis. Requirement for IMV increased with shorter duration of symptoms before hospital admission (adjusted OR 1.22 per day less, 95% CI 1.10-1.37, p<0.01, **<u>Table 1</u>**).

Seventy-nine percent (56/71) of all intubated patients required long IMV. Need for long IMV was associated with a short (≤1 day) duration from admission until intubation (20.0% (3/15) of patients with short IMV vs 53.6% (30/56) patients with long IMV, unadjusted OR (uOR) 4.6, 95% CI 1.17-18.16, p=0.02). Other factors associated with long IMV were transferral from other centers (28.6% (4/14) of patients with short IMV were transferred vs 69.6% (39/56) of patients with long IMV, uOR 5.73, 95% CI: 1.58-20.87, p<0.01), a higher total mean SOFA score during the second week after initial admission (8.4, 95% CI 5.83-10.9 in patients with short IMV vs 11.2, 95% CI 10.1-12.3, p<0.05 in patients with long IMV) as well as differences in SOFA score components (coagulation, hepatic impairment; for details see **<u>Supplementary table 1</u>**.)

IMV patients had a significantly higher risk of death from COVID-19 compared to non-IMV patients (1.1% (1/90) of non-IMV patients died vs 29.6% (21/71) of IMV patients, uOR 37.38, 95% CI 4.88-286.26, p<0.01). Sixty-three percent (44/70) of IMV patients underwent tracheotomy at a median time of 15 days (IQR 12-20) from intubation. Weaning from IMV was successfully concluded in 76.0% (38/50) of surviving IMV patients after a median of 42 days (IQR 16-66) from intubation. Eighteen percent (9/50) of patients remained dependent on intermittent IMV and 6.0% (3/50) on long-term oxygen therapy (LTOT) upon discharge or transferral.

Thirty-one percent (22/71) of all IMV patients required vvECMO (hereafter termed “ECMO”) treatment. ECMO was initiated a median of nine days (IQR 5.3-18.5) from intubation and continued for a median of 18 days (IQR 7.8-35.5). All ECMO patients required hemodialysis, 86.4% (19/22) underwent tracheotomy, 50% (11/22) had a VTE, and 50% (11/22) died. Seventy-five percent (53/71) of all patients with IMV and all (22/22) patients on ECMO received proning therapy. Details of the subgroup of patients receiving proning therapy have been reported elsewhere [32].

Hemodialysis was initiated in 30.4% (51/168) of patients, and in 66.2% (47/71) of patients with IMV. Hemodialysis was initiated after a median of eight days (IQR 4.5-14 days) following hospital admission and five days (IQR 2-8.8 days) after intubation. Thirty-eight percent (18/47) of IMV patients with hemodialysis died. For details on tracheotomy, ECMO and hemodialysis see **<u>Supplementary table 2</u>**.

As no evidence of efficacy of antiviral and anti-inflammatory treatments was available at the time, these were only used in a small subgroup of patients of this cohort (**<u>Supplementary table 3</u>**).

### Virological and routine laboratory data

Viral concentration data was available for 166/168 patients. On average, each patient had seven RT-PCR tests (sd: 5.3, min=1, max=29, including positive and negative result) from the day of symptom onset (or 10 days from first admission, if the date of symptom onset was not available) to the end of hospitalization, with tests performed every 8.4 days on average (sd: 8.8). Eighty-six patients had two final negative RT-PCR tests at the end of the disease course. Median first-measured viral concentration differed by 0.68 log_10_ viral copies between IMV and non-IMV patients (5.9, IQR 4.68-7.28 vs 5.22 log_10_ viral copies, IQR 4.49-7.28, respectively; p=0.12, **Figure 2A**), and median highest viral concentration by 1.19 log_10_ viral copies (6.7, IQR 5.35-7.62 vs 5.51 log_10_ viral copies, IQR 4.7-7.62, respectively; p=0.02, **Figure 2B**). Decline of viral concentration (**<u>Supplementary figure 1</u>**) was significantly slower in IMV versus non-IMV patients (–0.13, IQR –0.19-–0.08 vs –0.22 log_10_ viral concentration decrease / day, IQR –0.3-–0.08, respectively; p<0.01) (**Figure 2C, <u>Supplementary figure 2A-D</u>**). The duration of shedding was significantly longer in IMV patients than in non-IMV patients (median 33, IQR 26-46.75 vs 18 days, IQR 16-46.75, p=<0.01). We found no association between viral concentration and the duration from symptom onset to admission and no difference in first or in highest viral concentrations in patients requiring long versus short IMV (**<u>Supplementary figure 3A-D</u>, Supplementary table 1**).

**Figure 2:**
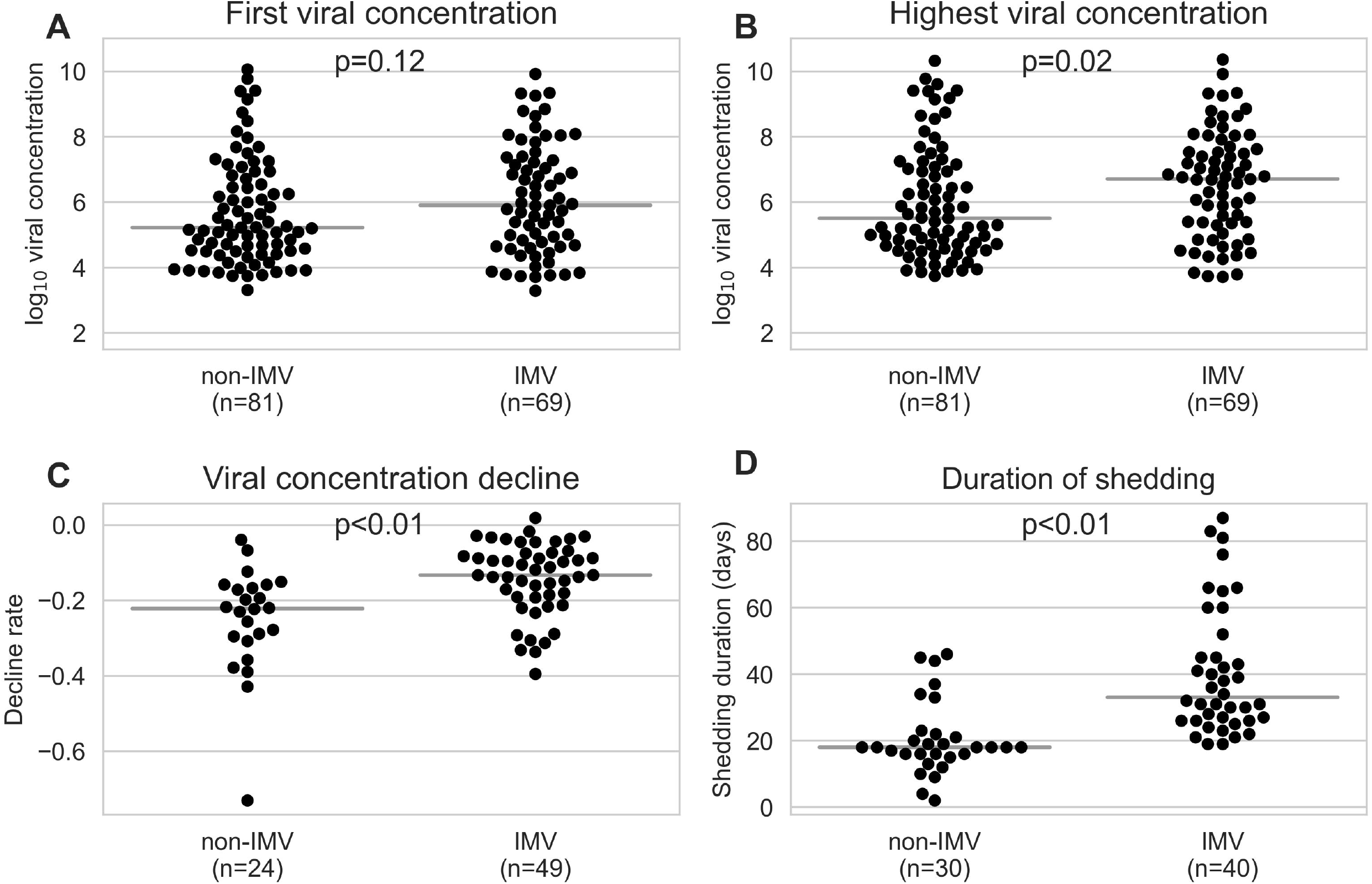
Comparison of viral concentration between patients with invasive mechanical ventilation (IMV) and those without IMV (non-IMV). A) First measured viral concentration: Median log_10_ viral concentration and (IQR) are 5.9 (4.68-7.28) for IMV patients and 5.22 (4.49-7.28) for non-IMV patients. B) Highest viral concentration: Median log_10_ viral concentrations and (IQR) are 6.7 (5.35-7.62) for IMV patients and 5.51 (4.7-7.62) for non-IMV patients. C) Differences in the slopes of log_10_ viral concentration decline rates were estimated using a linear regression of viral concentration from the full disease course of a patient and days since symptom onset (n=63) or admission (n=10) for patients with and without IMV. Only patients with at least four viral concentration measurements were included. D) Duration from symptom onset to the first of at least two final negative RT-PCR results for ventilated and non-ventilated patients. Median 33 days (IQR: 26-46.75) for IMV vs 18 days (IQR: 16-46.75) for non-IMV patients, p<0.01) Pairwise comparisons were performed using a Mann-Whitney *U* test. Grey horizontal lines indicate the median.

A statistically significant difference between non-IMV and IMV patients was observed in the levels of C-reactive protein (CRP), procalcitonin (PCT), Interleukin-6 (IL-6), lactate dehydrogenase, ferritin, white blood cell count, lymphocyte count, neutrophil-to-lymphocyte ratio, creatinine, urea, aspartate aminotransferase, creatine kinase, N-terminal prohormone of brain natriuretic peptide, and troponin **<u>(Supplementary table 4)</u>**. The course of 12 routine laboratory parameters over time in non-IMV and IMV patients is shown in Figure 3.

**Figure 3:**
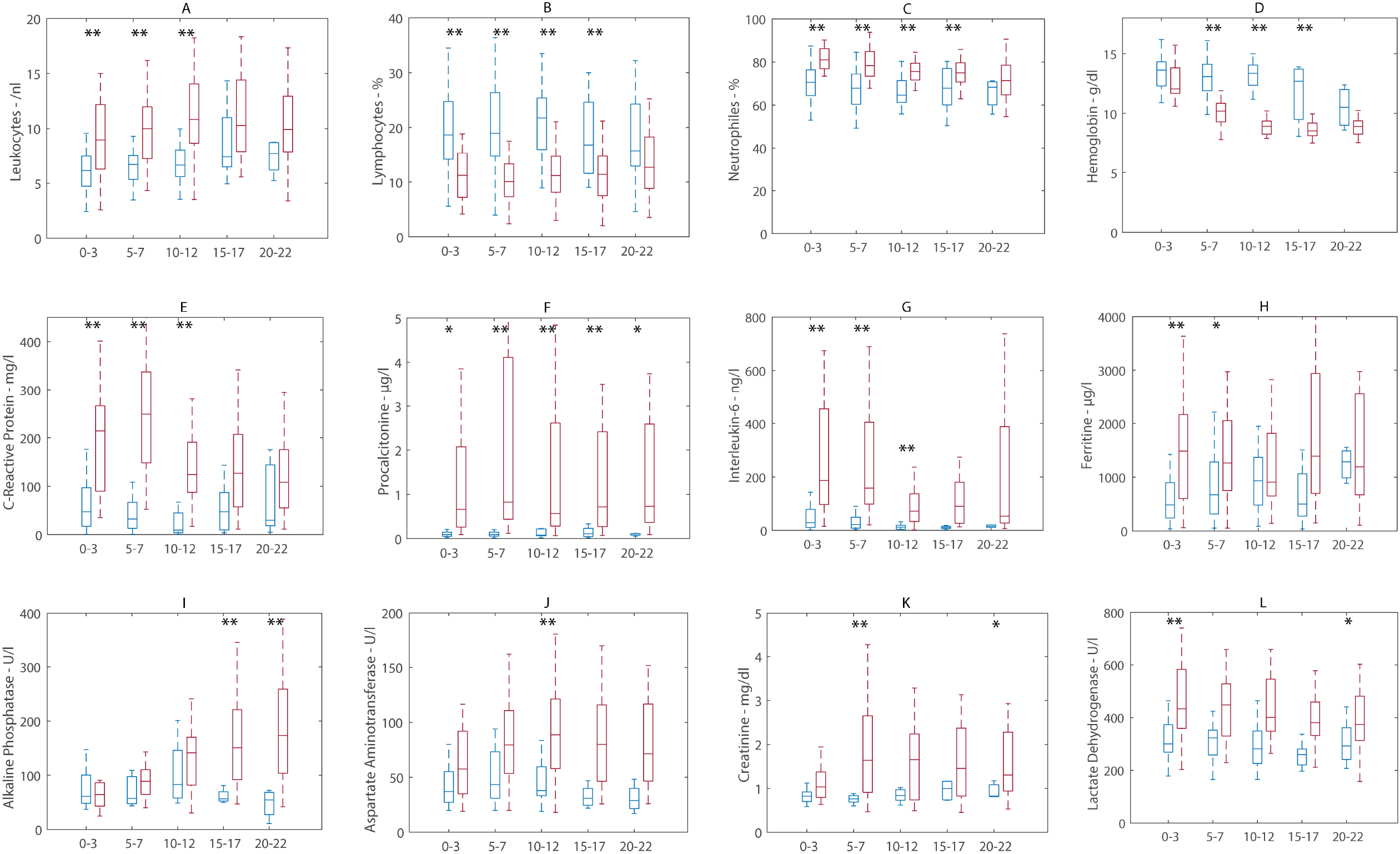
**A-L:** Comparison of laboratory parameters during the course of disease in IMV (red) versus non-IMV patients (blue). X-axis: days post admission. The boxes and lines are median 25th and 75th percentiles, Whiskers indicate the 1st and 99th percentile. A Welch’s t-test was used: *p<0.05, **p<0.01.

### Complications during treatment

Eighteen percent (31/168) of patients developed sepsis. Occurrence of sepsis was associated with organ replacement therapies, but not with patient-related risk factors (**<u>Supplementary table 5</u>**). The first sepsis episode occured after a median of 16 days (IQR 8-21) from hospital admission and 11.5 days (IQR 3-18.3 days) from intubation. In 15/31 (48.4%) of patients with sepsis, ECMO was initiated during or soon after occurrence of sepsis (median time from occurrence of sepsis and initiation of ECMO, 0 days, IQR 0-6.25 days).

Nineteen percent (32/168) of patients were diagnosed with at least one VTE. Due to evidence for increased risk of thromboembolic events in COVID-19 (36), therapeutic anticoagulation was introduced in all critically ill patients from April 2020. A quarter of VTEs (8/32) occurred without anticoagulatory treatment, 18.8% (6/32) under prophylactic anticoagulation, and 46% (15/32) under therapeutic anticoagulation. In 9.3% (3/32) of patients, VTE was diagnosed at autopsy and anticoagulation status at onset was unclear.

Twenty-four percent of patients (41/168) had at least one neurologic event during hospitalization, including hemorrhagic stroke (9/41, 22%), ischemic stroke (3/41, 7.3%), delirium (17/41, 41.5%), CIP/CIM (17/41, 41.5%), and epileptic seizure (3/41, 7.3%). Details of sepsis episodes, VTE and neurologic events are shown in **<u>Supplementary table 5</u>**.

### Outcome

Median time of hospital stay was 14 days (IQR 7-35) for all, 9 days (IQR 6-15.5) for non-IMV, and 49.5 days (IQR 36.8-82.5) for IMV patients.

Seventeen percent (29/168) of all patients died. Of all patients without therapy limitations (DNR/DNI), 13.6% (22/161) died, 8.7% (14/161) were transferred to other centers, and 77.6% (125/161) were discharged. Median time from first hospitalization until death was 33 days (IQR 16-98).

In univariate analyses, CCI ≥3 (uOR 4.35, 95% CI 1.52-12.45, p<0.01), short duration of symptoms (uOR 6.08, 95% CI 1.88-19.68, p<0.01), occurrence of sepsis (uOR 16.47, 95% CI 5.85-46.5, p<0.01), occurrence of VTE (uOR 7.58, 95% CI 2.88-19.97, p<0.01) and higher SOFA score during 1^st^ and 2^nd^ week after intubation were associated with death. There was a difference in the median first and highest viral concentration between survivors and non-survivors (6.84, IQR 4.99-7.91 vs 5.38, IQR 4.54-7.91, p=0.05; and 7.14, IQR 5.39-7.91 vs 5.86, IQR 4.77-7.91, p=0.04, respectively) (**<u>Supplementary table 6</u>**).

Of all discharged or transferred patients, 6.5% (9/139) still required IMV, 2.9% (4/139) LTOT, and 1.4% (2/139) new hemodialysis.

## Discussion

We analyzed a detailed clinical and virological dataset from a prospective observational cohort of COVID-19 patients hospitalized in a tertiary, ECMO/ARDS referral and weaning center in Germany during the first wave of the SARS-CoV-2 pandemic, before dexamethasone became standard of care. One of our major findings is that rapid clinical deterioration – as reflected by short duration of symptoms before hospital admission – is a highly relevant risk factor for both need of IMV in a multivariable risk model and death in univariate analyses in our cohort.

A shorter duration of symptoms was not associated with higher initial viral concentration. There was also no difference in initial viral concentration in more severely ill patients, i.e., those requiring IMV, compared to mildly ill patients. Yet, we found a significant increase of inflammatory markers such as CRP, PCT, and IL-6 at presentation and over time in IMV compared to non-IMV patients. Moreover, IMV patients had higher maximal concentrations, a slower decline as well as longer duration of shedding of SARS-CoV-2 compared to non-IMV patients. Higher maximal viral concentrations were also found in patients who died, compared to survivors.

Our findings underline that the early inflammatory host response to SARS-CoV-2 determines the course of disease more than pathogen factors such as initial SARS-CoV-2 RNA concentration [2, 39-41]. A short duration of symptoms before admission possibly reflects a rapid increase of the level of inflammation and might serve as an easy to assess risk factor for clinical deterioration [42]. During the further course of COVID-19, severe disease is characterized by the inability to rapidly reduce viral particles. A possible explanation for this phenotype – rapid deterioration after symptom onset, high inflammatory markers, and lack of efficient clearing of viral particles – might be the various kinds of immune dysregulation, as reported by our group and others [17, 43, 44].

Conflicting results have been published regarding the association between clinical severity and viral concentration for SARS-CoV-2 so far [33-38]. This could possibly be explained by a rather unbalanced proportion of mildly and severely affected patients in the respective studies as compared to our cohort [38]. However, it is undetermined whether higher maximal viral concentration and longer duration of shedding are a possible cause or an indicator of more severe organ damage and disease.

Regarding invasive treatment methods and complications, although it is difficult to separate cause from effect, onset of sepsis before or around the time of initiation of ECMO may indicate that sepsis drives clinical deterioration rather than ECMO driving sepsis, a topic requiring further investigation. In line with this, sepsis was a strong risk factor for death in our study.

Although a remarkable proportion (29.8%) of patients were transferred to our center due to severe ARDS, and overall 44.1% needed IMV, we report a comparatively low in-hospital mortality of 13.6% in patients without limitations of therapy. In comparison, Karagiannadis et al. reported in-hospital mortality of 22.2% [1] and Rieg et al. of 23.9% [45] in cohorts with 17.2% [1] and 32.9% [45] ventilated patients, respectively. Of note, our data show a high median length of hospital stay of 49.5 days for patients requiring IMV. By comparison, the median length of hospital stay for non-COVID ARDS patients was 17 days in a recent global multi-center prospective study [46]. There is growing evidence that the length of IMV-, ICU-, and inpatient treatment of patients with COVID-19 ARDS exceeds that of patients with ARDS unrelated to COVID-19 [42, 45].

Despite the long median duration of hospital stay, a considerable percentage of patients could not successfully be weaned off the ventilator (18%), and 6% required ongoing oxygen therapy following discharge. The mere number of deceased patients therefore depicts the burden of disease of COVID-19 only very incompletely, particularly with respect to long-term morbidity. The prospective approach of our study will allow us to evaluate long-term complications in the aftermath of COVID-19.

Prospective observational studies are often hampered by selection of patients with relatively mild disease courses due to need for informed consent. In our study, a deferred-consent model was applied, allowing for inclusion of critically ill patients. The high proportion of severely affected patients indicates that this selection bias does not apply to our data. On the other hand, one fifth of patients were only mildly affected and did not require oxygen therapy, representing a sub-cohort admitted for clinical observation or due to lacking the possibility of self-isolation.

In conclusion, we report a short duration of symptoms before clinical deterioration as a risk-factor for severe COVID-19, a finding that merits further exploration as an easily assessable prognostic factor. Second, we show that severely ill patients have higher maximal viral concentrations and a slower decline of viral concentration compared to mildly-affected patients. Third, our results demonstrate a comparatively long duration of inpatient treatment for patients with IMV, largely exceeding that described for non-COVID-19 ARDS patients. Our data give a comprehensive description of the clinical course, virological characteristics, organ support treatment, complications, and outcome of a representative cohort of patients in an unrestricted tertiary care healthcare setting with comparatively low mortality. The reported findings will be of value for both scientific progress in the fight against SARS-CoV-2 as well as for clinical management of critical care and weaning resources in the ongoing pandemic.

## Data Availability

there are no external dataset in this manuscript

## Supplementary figure legends

**Supplementary Figure 1**: Courses of viral concentration over time for A) IMV and B) non-IMV patients. Only patients with viral concentration measurements on at least four different days were included. If a patient had multiple viral concentration measurements on the same day, only the highest viral concentration was included for that day. If available, the first of at least two final negative PCRs is included with a viral concentration of 2.0 assigned. The x-axis displays the number of days since symptom onset, if available (n=63), or number of days since admission (n=10).

**Supplementary Figure 2**: Viral concentration decline rates, taking into account full and partial viral concentration courses for each patient. Decline rates were calculated using a linear regression for patients with at least four RT-PCR results. A, B) include positive RT-PCR results and the first of at least two final negative RT-PCR tests for a patient. C, D) only include positive RT-PCR test results. A, C) are based on all RT-PCR test results for a patient, while B, D) only take into account results from RT-PCR tests performed within 30 days of symptom onset (n=63), or date of admission (n=10) if date of symptom onset was unknown. Pairwise comparisons were performed using a Mann-Whitney *U* test. Grey horizontal lines indicate the median.

**Supplementary Figure 3**: Log_10_ viral concentration plotted against time in days from symptom onset to admission. A) and B) include data from invasive mechanically ventilated (IMV) patients, and C) and D) from non-IMV patients. A and C) show the first-measured viral concentration per patient, and B and D) illustrate the highest viral concentration per patient. Shaded areas indicate the 95% confidence interval. p – p-value, R – correlation coefficient.

