## Supplementary figures and images for "Clinical and Virological Characteristics of Hospitalized COVID-19 Patients in a German Tertiary Care Center during the First Wave of the SARS-CoV-2 Pandemic"

### Supplemental Figure 1

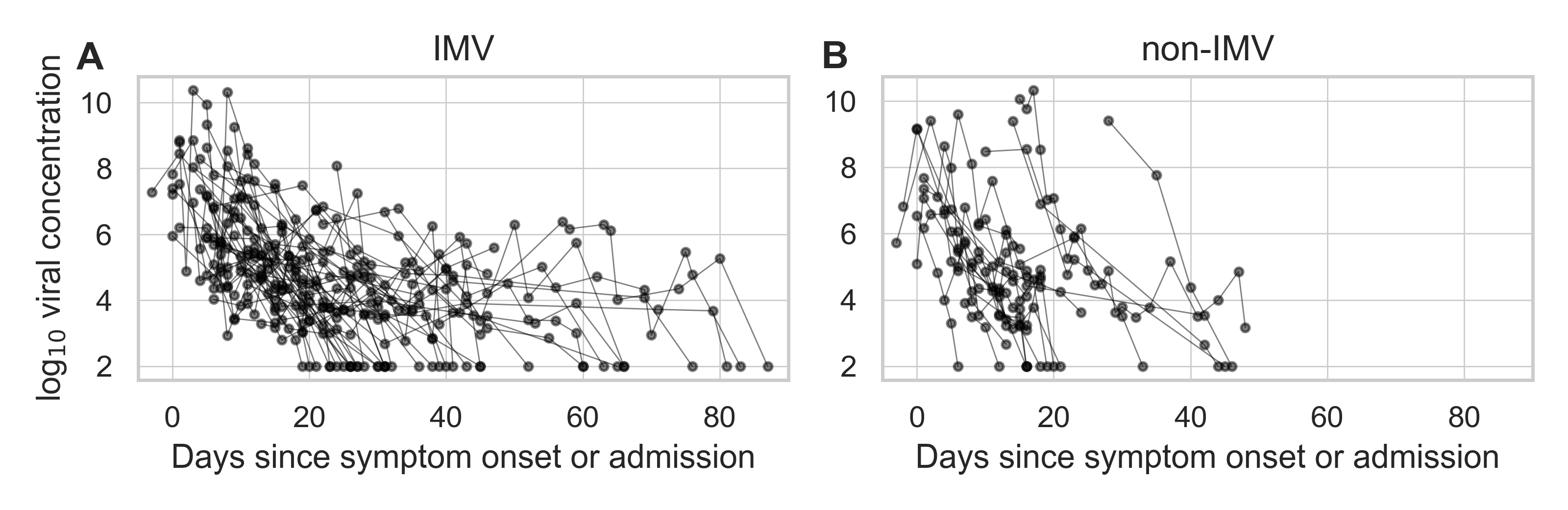

### Supplemental Figure 2

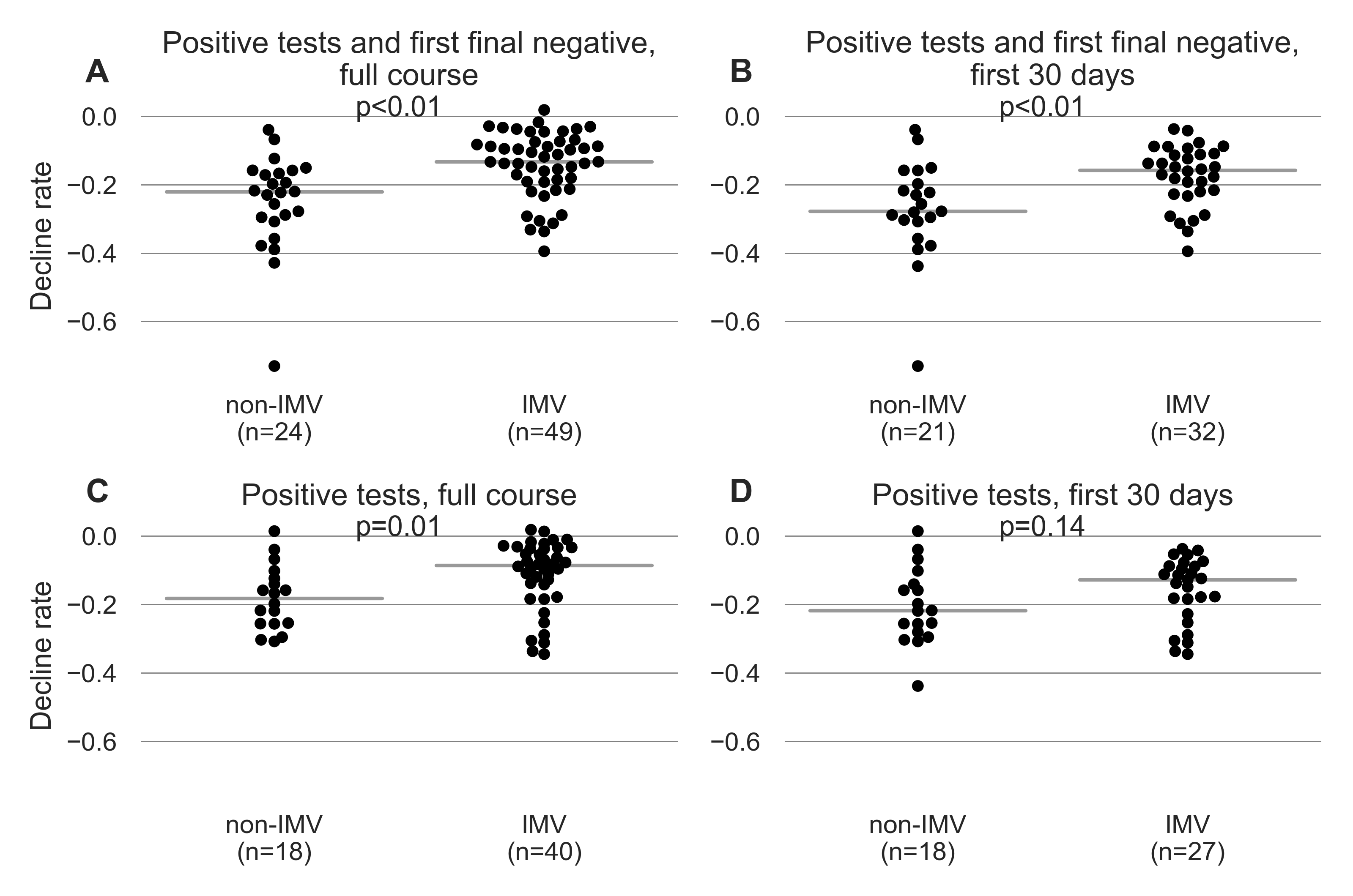
