## Supplemental Table 1 for "Clinical and Virological Characteristics of Hospitalized COVID-19 Patients in a German Tertiary Care Center during the First Wave of the SARS-CoV-2 Pandemic"

**Supplementary table 1:** **Short (<15 days) versus long (≥15 days) course of invasive mechanical ventilation**. Percentages and univariate analysis of associated patient factors, SOFA score, and SOFA score components in ventilated patients. Bold indicates statistical significance. IMV – invasive mechanical ventilation, uOR – unadjusted Odds Ratio, CI – confidence interval, BMI – body mass index, CCI – Charlson comorbidity index, SOFA – sequential organ failure assessment. Differences in viral concentrations are based on the highest viral concentration measurement for each patient.

|  | **Short IMV**  **percentage, n/available N** | **Long IMV**  **percentage n/available N** | **uOR (95% CI), Chi-square test** | **p-value** |
| --- | --- | --- | --- | --- |
| Total | 21.1%, 15/71 | 78.9%, 56/71 |  |  |
| 18-59 years | 30.0%, 5/15 | 42.9%, 24/56 | Reference | Reference |
| 60-69 years | 20.0%, 3/15 | 33.9%, 19/56 | 1.31 (0.28-6.23) | 0.73 |
| 70-79 years | 20.0%, 3/15 | 19.6%, 11/56 | 0.76 (0.15-3.78) | 0.74 |
| ≥ 80 years | 26.7%, 4/15 | 3.6%, 2/56 | 0.10 (0.01-0.73) | 0.017 |
| Male gender | 73.3%, 11/15 | 69.6%, 39/56 | 0.83 (0.23-3.00) | 0.78 |
| BMI ≥ 30 kg/m^2^ | 28.6%, 4/14 | 41.8%, 23/55 | 1.80 (0.50-6.45) | 0.36 |
| CCI ≥3 | 73.3%, 11/15 | 55.4%, 31/56 | 0.45 (0.13-1.60) | 0.21 |
| Cardiovascular disease | 73.3%, 11/15 | 64.3%, 36/56 | 0.65 (0.18-2.32) | 0.51 |
| Diabetes | 20.0%, 3/15 | 25.0%, 14/56 | 1.33 (0.33-5.42) | 0.69 |
| Chronic pulmonary disease | 20.0%, 3/15 | 21.4%, 12/56 | 1.09 (0.26-4.50) | 0.90 |
| Chronic kidney disease | 6.7%, 1/15 | 8.9%, 5/56 | 1.37 (0.15-12.72) | 0.77 |
| Chronic neurological disease | 6.7%, 1/15 | 10.7%, 6/56 | 1.68 (0.19-15.14) | 0.64 |
| Days between symptom onset and admission  ≤ 5 days, available n/N | 54.5%, 6/11 | 43.2%, 19/44 | 1.10 (0.29-4.14) | 0.89 |
| Days from admission until intubation  ≤ 1 day | 20.0%, 3/15 | 53.6%, 30/56 | **4.60 (1.17-18.16)** | **0.02** |
| Patients transferred from other centers | 28.6%, 4/14 | 69.6%, 39/56 | **5.73 (1.58-20.87)** | **<0.01** |
|  | **mean (sd) (95% CI), available n/N** | **mean (sd) (95% CI)**,  **available n/N** | **Difference of means**  **(95% CI)**  **(Welch’s t-test)** | **p-value** |
| SOFA total score max. 1st week after hospital admission | 9.44 (2.92) (7.20-11.7), 9/15 | 11.5 (2.97) (10.1-12.8), 22/56 | 2.01 (-0.46-4.48) | 0.10 |
| SOFA total score max. Score 2^nd^ week after hospital admission | 8.40 (3.53) (5.87-10.9), 10/15 | 11.2 (2.92) (10.1-12.3), 30/56 | 2.8 (0.13-5.47) | **0.04** |
| SOFA total score max. | 10.8 (1.87) (9.46-12.1), 10/15 | 12.7 (2.64) (11.8-13.6), 38/56 | 1.88 (0.36-3.41) | **0.02** |
| SOFA respiratory component max. | 3.20 (0.63) (2.75-3.65), 10/15 | 3.32 (0.62) (3.11-3.52), 38/56 | 0.12 (-0.37;0.60) | 0.61 |
| SOFA coagulation component max. | 0.30 (0.48) (-0.05-0.65)  10/15 | 1.03 (1.05) (0.68-1.37)  38/56 | 0.73 (0.26-1.19) | **<0.01** |
| SOFA hepatic component max. | 0.40 (0.70) (-0.10-0.90)  10/15 | 1.55 (1.37) (1.10-2.00)  38/56 | 1.15 (0.51-1.79) | **<0.01** |
| SOFA cardiovascular component max. | 3.50 (0.71) (2.99-4.01), 10/15 | 3.58 (0.86) (3.30-3.86), 38/56 | 0.08 (-0.48-0.64) | 0.77 |
| SOFA neurological component. | 4.00 (0.00) (4.00-4.00)  10/15 | 3.84 (0.59) (3.65-4.04)  38/56 | -0.16 (-0.35-0.04) | 0.11 |
| SOFA renal component max. | 1.40 (1.58) (0.27-2.53)  10/15 | 2.42 (1.41) (1.96-2.88)  38/56 | 1.02 (-0.16-2.21) | 0.09 |
|  | **Median (IQR), available n/N** | **Median (IQR), available n/N** | **Difference of medians (IQR of pairwise differences), Mann-Whitney *U* test** | **p-value** |
| Highest viral concentration | 7.19  (5.74-7.46), 15/15 | 6.63  (5.31-7.46),  54/56 | 0.56 (–2.42-1.04) | 0.19 |
| First viral concentration | 5.69 (4.83-7.11), 15/15 | 5.92 (4.71-7.11), 54/56 | 0.24 (–2.17-1.40) | 0.54 |
