## Supplemental Table 2 for "Clinical and Virological Characteristics of Hospitalized COVID-19 Patients in a German Tertiary Care Center during the First Wave of the SARS-CoV-2 Pandemic"

**Supplementary table 2**: **Tracheotomy, vvECMO and hemodialysis**. Percentages and univariate analysis of associated patient factors in ventilated patients. Bold indicates statistical significance. vvECMO – veno-venous extra corporeal membrane oxygenation, uOR – unadjusted Odds Ratio, CI – confidence interval, BMI – body mass index, CCI – Charlson comorbidity index.

|  | **Patients with tracheotomy** | | | **Patients on vvECMO** | | | **Patients on hemodialysis** | | |
| --- | --- | --- | --- | --- | --- | --- | --- | --- | --- |
|  | **percentage, n/ available N** | **uOR (95% CI)** | **p-**  **value**  **chi-**  **square** | **percentage, n/ available N** | **uOR (95% CI)** | **p-**  **value**  **chi-**  **square** | **percentage, n/ available N** | **uOR (95% CI)** | **p-**  **value**  **chi-**  **square** |
| Total | 62.9%, 44/70 |  |  | 31.0%, 22/71 |  |  | 66.2%, 47/71 |  |  |
| 18-59 years | 18.2%, 8/44 | reference |  | 63.6%, 14/22 | reference |  | 31.9%, 15/47 | reference |  |
| 60-69 years | 29.5%, 13/44 | 0.99 (0.32-3.16) | 0.99 | 18.2%, 4/22 | **0.29**  **(0.07-0.88)** | **0.02** | 36.2%, 17/47 | 3.17  (0.92-10.91) | 0.06 |
| 70-79 years | 25.0%, 11/44 | 2.24 (0.51-9.84) | 0.28 | 18.2%, 4/22 | 0.43  (0.11-1.69) | 0.21 | 23.4%, 11/47 | 3.42  (0.79-14.89) | 0.08 |
| ≥ 80 years | 4.5%, 2/44 | 0.31 (0.05-1.95) | 0.20 | 0/22 |  |  | 8.5%, 4/47 | 1.87  (0.29-11.84) | 0.50 |
| Male gender | 72.7%, 32/44 | 1.19 (0.41-3.44) | 0.75 | 77.3%, 17/22 | 1.65  (0.52-5.27) | 0.39 | 78.7%, 37/47 | **3.13**  **(1.08-9.08)** | **0.03** |
| BMI ≥ 30 kg/m^2^ | 36.4%, 16/44 | 0.75 (0.28-2.06) | 0.58 | 31.8%, 7/22 | 0.63  (0.22-1.83) | 0.39 | 42.6%, 20/47 | 1.59  (0.55-4.62) | 0.40 |
| CCI ≥3 | 56.8%, 25/44 | 0.82 (0.31-2.21) | 0.70 | 40.9%, 9/22 | **0.34**  **(0.12-0.95)** | **0.04** | 70.2%, 33/47 | **3.93**  **(1.39-11.07)** | **<0.01** |
| Cardiovascular disease | 61.4%, 27/44 | 0.59 (0.20-1.70) | 0.32 | 59.1%, 13/22 | **0.64**  **(0.22-1.81)** | **0.04** | 74.5%, 35/47 | **2.92**  **(1.04-8.21)** | **0.04** |
| Diabetes | 27.3%, 12/44 | 1.57 (0.48-5.12) | 0.45 | 22.7%, 5/22 | 0.91  (0.28-2.98) | 0.87 | 27.7%, 13/47 | 1.91  (0.55-6.67) | 0.29 |
| Chronic pulmonary disease | 18.2%, 8/44 | 0.74 (0.23-2.44) | 0.62 | 18.2%, 4/22 | 0.77  (0.21-2.75) | 0.69 | 27.7%, 13/47 | 4.21  (0.86-20.47) | 0.06 |
| Chronic kidney disease | 11.4%, 5/44 | 3.20  (0.35-29.07) | 0.28 | 9.1%, 2/22 | 1.13  (0.19-6.65) | 0.90 | 14.6%, 6/41 | Not calculable | **0.02** |
| Chronic neurological disease | 6.8%, 3/44 | 0.56 (0.10-3.00) | 0.50 | 4.8%, 1/21 | **0.29**  **(0.03-2.53)** | **0.02** | 10.8%, 4/37 | 0.85  (0.17-4.18) | 0.84 |
| Days between symptom onset and admission ≤ 5 days | 64.9%, 24/37 | 2.03 (0.68-6.04) | 0.20 | 65.0%, 13/20 | **3.37 (1.24-9.17)** | **0.01** | 60.0%, 24/40 | **3.33 (1.53-7.26)** | **<0.01** |
