## Supplemental Table 3 for "Clinical and Virological Characteristics of Hospitalized COVID-19 Patients in a German Tertiary Care Center during the First Wave of the SARS-CoV-2 Pandemic"

**Supplementary table 3**: **Antiviral, disease modifying, and immunosuppressant drugs and biologics**. Overview of percentages of treated patients, dose and indication used in this cohort. p.o. – per os, i.v. – intravenously, ARDS – acute respiratory distress syndrome, COPD – chronic obstructive pulmonary disease. SARS-CoV-2 - Severe Acute Respiratory Syndrome Coronavirus 2.

| **Drug** | **Percentage, n/N  treated patients** | **Daily dose and route of administration** | **Indication** |
| --- | --- | --- | --- |
| Prednisolon equivalent ≥40mg/d | 10.1%, 17/168 | 250 mg i.v. | Allergic reaction (n = 1)  Glottis edema (n = 1) |
|  |  | 100 mg i.v. | Fibroproliferative ARDS (n = 2)  Cryptogenic organizing pneumonia (n = 1)  Allergic reaction (n = 1)  Obstruction (n = 1)  Stridor post extubation (n = 1)  Not evaluable (n = 1) |
|  |  | 80 mg  i.v. | Not evaluable (n = 2) |
|  |  | 50 mg i.v. | Fibroproliferative ARDS (n = 2)  Giant cell arteritis (n = 1)  Obstruction (n = 1) |
|  |  | 40 mg i.v. | Acute exacerbation of COPD (n = 2) |
| Dexamethason | 9.5%, 16/168 | 6 mg p.o. | SARS-CoV-2 (n = 13) |
|  |  | 20 mg i.v. | Suspected hemophagocytic lymphohistiocytosis (n = 1) |
|  |  | 22 mg i.v. | Suspected hemophagocytic lymphohistiocytosis (n = 2) |
| Remdesivir | 3.6%, 6/168 | 100-200 mg p.o. | SARS-CoV-2 |
| Lopinavir/Ritonavir |  | 200/50 mg p.o. | HIV-Infection (n = 1) |
|  |  | 800/200mg p.o. | SARS-CoV-2 (n = 1) |
| Hydroxychloroquine | 1.2%, 2/168 | 400mg p.o. | Systemic lupus erythematosus (n = 1) |
|  |  | 400mg p.o. | SARS-CoV-2 (n = 1) |
| Tocilizumab | 0.6%, 1/168 | 162mg i.v. | SARS-CoV-2 |
| Anakinra | 1.8%, 3/168 | 200 mg i.v. | Suspected hemophagocytic lymphohistiocytosis  (n = 1) |
|  |  | 100 mg  i.v. | Suspected hemophagocytic lymphohistiocytosis  (n = 1) |
|  |  | 200 mg s.c. | Suspected hemophagocytic lymphohistiocytosis  (n = 1) |
| Immunoglobulins | 1.8%, 3/168 | 10 g i.v.  (cumulative dose 40g i.v.) | Suspected hemophagocytic lymphohistiocytosis  (n = 2) |
|  |  | 10g i.v.  (cumulative dose 50 g i.v.) | Suspected hemophagocytic lymphohistiocytosis  (n = 1) |
