## Supplemental Table 4 for "Clinical and Virological Characteristics of Hospitalized COVID-19 Patients in a German Tertiary Care Center during the First Wave of the SARS-CoV-2 Pandemic"

**Supplementary table 4**: **Laboratory parameters from first 72h after admission** in all, non-IMV and IMV patients. Data from all available n/N are shown as medians and IQRs. Bold indicates statistical significance.

IMV – invasive mechanical ventilation, CRP – C-reactive protein, PCT – Procalcitonin, LDH – lactate dehydrogenase, IL-6 – Interleukin 6, WBC – white blood cell count, NLR – neutrophil-to-lymphocyte-ratio, Hb – Hemoglobin, ALAT – alanine aminotransferase, ASAT – aspartate aminotransferase, CK – creatine kinase, NT-proBNP – N-terminal prohormone of brain natriuretic peptide.

|  | **All** | **Non-IMV** | **IMV** |  |
| --- | --- | --- | --- | --- |
| **Parameter** | **median (IQR), available n/N** | **median (IQR)**  **available n/N** | **median (IQR)**  **available n/N** | **p-value**  **Mann-Whitney *U* test** |
| CRP mg/L | 68.2  (25.1-124.1),  117/168 | 50.8  (14.7-103.1)  77/90 | 138  (68.2-372.7)  35/71 | **<0.01** |
| PCT µg/L | 0.1  (0.06-0.25)  114/168 | 0.08  (0.05-0.13)  74/90 | 0.27  (0.11-0.85),  35/71 | **<0.01** |
| LDH U/L | 341  (285-460)  114/168 | 303  (263-392)  75/90 | 469  (360.5-622.8),  34/71 | **<0.01** |
| IL-6 ng/L | 48.7  (18-118.6)  82/168 | 27.1  (10.7-72.7),  59/90 | 151.5  (79.8-151.5),  20/71 | **<0.01** |
| Ferritin µg/L | 651.8  (311.4-1640.8)  89/168 | 543.7  (244-1008.4)  57/90 | 1384.5  (524.9-2063.9),  29/71 | **<0.01** |
| WBC /nL | 6.5  (4.9-9.2)  117/168 | 6.04  (3.4-7.8)  77/90 | 7.86  (6-11.7), 35/71 | **<0.01** |
| Lymphocytes /nL | 0.9  (0.65-1.32)  114/168 | 1.01  (0.7-1.4),  75/90 | 0.8  (0.6-1), 34/71 | **0.02** |
| Neutrophils /nL | 4.6  (3.1-6.3)  105/168 | 4.2  (2.7-5.8),  74/90 | 5.57  (4.1-8.9), 27/71 | **<0.01** |
| NLR | 4.6  (3.0-7.0)  105/168 | 4.18  (2.7-5.8),  74/90 | 6.6  (4.2-10.721), 27/71 | **<0.01** |
| Hb mg/dl | 13.3  (11.6-14.4),  117/168 | 13.6  (11.9-14.5),  77/90 | 12.7  (11.2-13.9), 35/71 | 0.08 |
| Platelets /nL | 198  (142-266.5)  117/168 | 201  (151.5-277.5),  77/90 | 194  (139-253), 35/71 | 0.68 |
| Creatinine mg/dL | 0.9  (0.8-1.3),  117/168 | 0.88  (0.8-1.1),  77/90 | 1.17  (0.9-1.6), 35/71 | **<0.01** |
| Urea mg/dL | 33.5  (22-51.8),  100/168 | 26  (19-39.5),  62/90 | 41  (31-75.5), 33/71 | **<0.01** |
| Lactate mg/dL | 12  (9-15), 99/168 | 12  (8-14), 59/90 | 13  (10-15), 35/71 | 0.10 |
| Total bilirubin mg/dL | 0.47  (0.31-0.69), 116/168 | 0.45  (0.27-0.65), 76/90 | 0.52  (0.37-0.77), 35/71 | 0.09 |
| ALAT U/L | 31  (20-45), 117/168 | 32  (19-46), 77/90 | 30  (20-43), 35/71 | 0.92 |
| ASAT U/L | 43  (30.5-70.5),  97/168 | 38  (28-55),  59/90 | 59  (40.5-91.6), 33/71 | **<0.01** |
| CK U/L | 95  (56.5-241),  101/168 | 83  (56-132), 67/90 | 260.5  (82.3-700.5), 30/71 | **<0.01** |
| NT-proBNP pg/mL | 202.5  (86-951.3),  106/168 | 136  (42-406), 69/90 | 672.5  (193.8-1815.3), 32/71 | **<0.01** |
| Troponin T ng/L | 12.5  (6-35.5),  74/168 | 7  (5.5-19), 49/90 | 23  (10.5-66), 21/71 | **<0.01** |
| D-Dimers mg/L | 1.8  (0.9-3.2),  43/168 | 0.97  (0.6-2.1),  13/90 | 2.17  (1-3.7), 27/71 | **0.02** |
