## Supplemental Table 5 for "Clinical and Virological Characteristics of Hospitalized COVID-19 Patients in a German Tertiary Care Center during the First Wave of the SARS-CoV-2 Pandemic"

**Supplementary table 5: Sepsis, venous thromboembolic and neurologic events.** Percentages and univariate analysis of associated patient factors. Bold indicates statistical significance.

uOR – unadjusted Odds Ratio, CI – confidence interval, BMI – body mass index, CCI – Charlson comorbidity index, IMV – invasive mechanical ventilation, vvECMO – veno-venous extracorporeal membrane oxygenation.

|  | **Patients with sepsis** | | | **Patients with venous thromboembolic event** | | | **Patients with neurologic event** | | |
| --- | --- | --- | --- | --- | --- | --- | --- | --- | --- |
|  | **percentage, n/ available N** | **uOR (95% CI)** | **p-**  **value**  **chi-**  **square** | **percentage, n/ available N** | **uOR (95% CI)** | **p-**  **value**  **chi-**  **square** | **percentage, n/ available N** | **uOR (95% CI)** | **p-**  **value**  **chi-**  **square** |
| Total | 18.5%, 31/168 |  |  | 19.0%, 32/168 |  |  | 24.4%, 41/168 |  |  |
| 18-59 years | 35.5%, 11/31 | reference |  | 46.8%, 15/32 | reference |  | 31.7%, 13/41 | reference |  |
| 60-69 years | 29.0%, 9/31 | 2.03 (0.75-5.5) | 0.16 | 25.0%, 8/32 | 1.2 (0.46-3.16) | 0.71 | 29.3%, 12/41 | 2.5 (1.0-6.2) | 0.05 |
| 70-79 years | 22.6%, 7/31 | 1.52 (0.53-4.33) | 0.43 | 21.9%, 7/32 | 1.05 (0.39-2.86) | 0.92 | 29.3%, 12/41 | 2.6 (1.0-6.5) | 0.04 |
| ≥ 80 years | 12.9% 4/31 | 1.60 (0.45-5.80) | 0.46 | 6.2%, 2/32 | 0.49 (0.10-2.38) | 0.37 | 9.8%, 4/41 | 1.33 (0.38-4.67) | 0.65 |
| Male gender | 77.4%, 24/31 | 1.97  (0.79-4.89) | 0.12 | 71.9%, 23/32 | 1.39 (0.60-3.25) | 0.44 | 63.4%, 26/41 | 0.85 (0.41-1.79) | 0.68 |
| BMI ≥ 30 kg/m^2^ | 29.0%, 9/31 | 0.78 (0.33-1.84) | 0.56 | 37.5%, 12/32 | 1.34 (0.60-3.03) | 0.47 | 40.0%, 16/40 | 1.49 (0.70-3.12) | 0.30 |
| CCI ≥3 | 64.5%, 20/31 | 2.01 (0.89-4.52) | 0.08 | 53.1%, 17/32 | 1.13 (0.52-2.45) | 0.75 | 70.7%, 29/41 | **3.06 (1.43-6.54)** | **<0.01** |
| Cardiovascular disease | 67.7%, 21/31 | 1.73 (0.76-3.96) | 0.18 | 56.2%, 18/32 | 0.95 (0.44-2.08) | 0.91 | 77.5%, 31/40 | **2.96 (1.34-6.54)** | **<0.01** |
| Diabetes | 25.8%, 8/31 | 1.55 (0.62-3.89) | 0.35 | 21.8%, 7/32 | 1.18 (0.46-3.04) | 0.72 | 26.8%, 11/41 | 1.75 (0.77-4.01) | 0.18 |
| Chronic pulmonary disease | 16.1%, 5/31 | 0.95 (0.33-2.74) | 0.92 | 18.6%, 6/32 | 1.20 (0.44-3.24) | 0.72 | 29.3%, 12/41 | **2.88 (1.22-6.74)** | **0.01** |
| Chronic kidney disease | 6.5% 2/31 | 0.38 (0.08-1.72) | 0.16 | 12.5%, 4/32 | 0.88 (0.28-2.80) | 0.83 | 14.6%, 6/41 | 1.11 (0.41-3.03) | 0.84 |
| Chronic neurological disease | 12.9%, 4/31 | 1.54 (0.46-5.15) | 0.49 | 15.6%, 5/32 | 2.10 (0.68-6.56) | 0.19 | 12.2%, 5/41 | 1.46 (0.48-4.50) | 0.50 |
| Days between symptom onset and admission ≤ 5 days | 59.3%, 16/27 | **2.7 (1.13-6.45)** | **0.02** | 61.5%, 16/26 | **2.53 (1.05-6.09)** | **0.03** | 63.3%, 19/30 | **3.12 (1.36-7.20)** | **<0.01** |
| IMV | 100%, 31/31 | n/a | **<0.01** | 87.5%, 28/32 | **14.62 (4.84-44.30)** | **<0.01** | 90.2%, 37/41 | **18.2 (6.61-50.06)** | **<0.01** |
| Long IMV | 87.1%, 27/31 | 2.47 (0.7-8.69) | 0.14 | 89.2%, 25/28 | 3.13 (0.80-12.29) | 0.09 | 83.3%, 30/36 | 1.67 (0.52-5.30) | 0.38 |
| Late tracheotomy | 52.4%, 11/21 | 0.93 (0.29-3.01) | 0.9 | 57.1%, 12/21 | 1.33 (0.41-4.33) | 0.63 | 41.7%, 10/24 | 0.35 (0.11-1.21) | 0.09 |
| Hemo-  dialysis | 83.9%, 26/31 | **23.29 (8.14-66)** | **<0.01** | 68.5%, 22/32 | **8.12 (3.47-19.04)** | **<0.01** | 61.0%, 25/41 | **6.07 (2.83-12.10)** | **<0.01** |
| vvECMO | 48.4% 15/31 | **17.41 (6.17-49.09)** | **<0.01** | 34.4%, 11/32 | **5.96 (2.30-15.47)** | **<0.01** | 19.5%, 8/41 | 1.96 (0.76-5.07) | 0.16 |
