## Supplemental Table 6 for "Clinical and Virological Characteristics of Hospitalized COVID-19 Patients in a German Tertiary Care Center during the First Wave of the SARS-CoV-2 Pandemic"

**Supplementary table 6: Comparison of surviors and non-survivors.** Percentages and univariate analysis of associated patient factors, SOFA score and SOFA score components. Bold indicates statistical significance. uOR – unadjusted Odds Ratio, CI – confidence interval, IQR- interquartile range,  BMI – body mass index, CCI – Charlson comorbidity index, VTE- venous thromboembolic event, SOFA – sequential organ failure assessment.

|  | **Survivors**  **percentage,**  **n/ available N** | **Non-Survivors**  **percentage,**  **n/ available N** | **uOR (95% CI)** | **p-value**  **chi-square test** |
| --- | --- | --- | --- | --- |
| Total | 86.3%, 139/161 | 13.7%, 22/161 |  |  |
| Age in years, median (IQR) | 60 (45-71) | 66.5 (53.75-74.5) |  |  |
| 18-59 years | 49.6%, 69/139 | 40.9%, 9/22 | Reference | Reference |
| 60-69 years | 23.0%, 32/139 | 18.2%, 4/22 | 0.96 (0.27-3.35) | 0.94 |
| 70-79 years | 20.1%, 28/139 | 22.7%, 5/22 | 1.36 (0.42-4.45) | 0.60 |
| ≥ 80 years | 7.2%, 10/139 | 18.2%, 4/22 | 3.06 (0.79-11.85) | 0.09 |
| Female | 36.7%, 51/139 | 22.7%, 5/22 | Reference |  |
| Male | 63.3% (88/139) | 77.3%, 17/22 | 1.97 (0.69-5.66) | 1.97 |
| BMI ≥ 30 kg/m^2^ | 32.8% 42/128 | 36.4%, 8/22 | 1.17 (0.46-3.00) | 0.74 |
| CCI ≥3 | 43.9% 61/139 | 77.3%, 17/22 | **4.35 (1.52-12.45)** | **<0.01** |
| Cardiovascular disease | 51.8%, 72/161 | 77.3%, 17/22 | **3.16 (1.10-9.05)** | **0.03** |
| Diabetes | 18.0%, 25/139 | 18.2%, 4/22 | 1.01 (0.32-3.25) | 0.98 |
| Chronic pulmonary disease | 15.8%, 22/139 | 22.7%, 5/22 | 1.56 (0.52-4.68) | 0.42 |
| Chronic kidney disease | 13.0%, 18/139 | 9.1%, 2/22 | 0.67 (1.45-3.12) | 0.61 |
| Chronic neurological disease | 7.9%, 11/139 | 9.1%, 2/22 | 1.16 (0.24-5.64) | 0.85 |
| Days between symptom onset and admission  ≤ 5 days, | 36.5%, 42/115 | 75.0%, 12/16 | **5.53 (1.67-18.32)** | **<0.01** |
| Primary hospitalization COVID-19 or transferal, n | 132 | 21 |  |  |
| Transfer from other centers | 28.0%, 37/132 | 47.7%, 10/22 | 2.33 (0.91-5.96) | 0.07 |
| Sepsis | 11.5%, 16/139 | 68.2%, 15/22 | **16.47 (5.84-46.5** | **<0.01** |
| VTE | 13.7%, 19/139 | 54.5%, 12/22 | 7.58 (2.88-19.97) | **<0.01** |
| Neurologic complication | 24.4%, 34/139 | 31.8%, 7/22 | 1.44 (0.54-3.83) | 0.46 |
|  | **mean (sd) (95% CI), available n/N** | **mean (sd) (95% CI) , available n/N** | **Difference of means**  **(95% CI) (Welch’s t-test)** |  |
| SOFA total score max. 1st week after hospital admission | 7.75 (4.51) (6.53-8.97), 55/139 | 11.4 (4.30) (8.32-14.5),  15/22 | 3.65 (0.43-6.88) | **0.03** |
| SOFA total score max Score 2^nd^ week after hospital admission | 9.08 (4.23) (7.88-10.3),  50/139 | 12.5 (3.55) (10.3-14.6),  13/22 | 3.38 (0.99-5.77) | **0.01** |
| SOFA total score max. | 9.08 (4.86) (7.98-10.2)  77/139 | 15.9 (2.72) (14.4-17.4)  15/22 | 6.79 (4.97-8.61) | **<0.01** |
| SOFA respiratory component max. | 2.70 (1.29) (2.41-2.99),  77/139 | 3.87 (0.35) (3.67-4.06)  15/22 | 1.17 (0.82-1.51) | **<0.01** |
| SOFA coagulation component max. | 0.64 (0.90) (0.43-0.84),  77/139 | 2.00 (1.31) (1.27-2.73),  15/22 | 1.36 (0.62-2.11) | **<0.01** |
| SOFA hepatic component max. | 0.82 (1.23) (0.54-1.10)  77/139 | 2.40 (1.40) (1.62-3.18)  15/22 | 1.58 (0.77-2.40) | **<0.01** |
| SOFA cardiovascular component max. | 2.45 (1.70) (2.07-2.84)  77/139 | 4.00 (0.00) (4.00-4.00)  15/22 | 1.55 (1.16-1.93) | **<0.01** |
| SOFA neurological component max. | 2.64 (1.78) (2.23-3.04)  77/139 | 4.00 (0.00) (4.00-4.00)  15/22 | 1.36 (0.96-1.77) | **<0.01** |
| SOFA renal component max. | 1.79 (1.58) (1.43-2.15)  77/139 | 2.73 (1.33) (1.99-3.47)  15/22 | 0.94 (0.14-1.75) | **0.02** |
|  | **Median (IQR), available n/N** | **Median (IQR), available n/N** | **Difference of medians (IQR of pairwise differences), Mann-Whitney *U* test** |  |
| First viral concentration | 5.38 (4.54-7.91), 130/151 | 6.84 (4.99-7.91), 21/151 | 1.47 (–0.83-2.68) | 0.05 |
| Highest viral concentration | 5.86 (4.77-7.91), 130/151 | 7.14 (5.39-7.91), 21/151 | 1.28 (–0.79-2.55) | **0.04** |
